# Applications of Large Language Models in Psychiatry: A Systematic Review

**DOI:** 10.1101/2024.03.28.24305027

**Authors:** Mahmud Omar, Shelly Soffer, Alexander W Charney, Isotta Landi, Girish N Nadkarni, Eyal Klang

## Abstract

**Background:** With their unmatched ability to interpret and engage with human language and context, large language models (LLMs) hint at the potential to bridge AI and human cognitive processes. This review explores the current application of LLMs, such as ChatGPT, in the field of psychiatry.

**Methods:** We followed PRISMA guidelines and searched through PubMed, Embase, Web of Science, and Scopus, up until March 2024.

**Results:** From 771 retrieved articles, we included 16 that directly examine LLMs’ use in psychiatry. LLMs, particularly ChatGPT and GPT-4, showed diverse applications in clinical reasoning, social media, and education within psychiatry. They can assist in diagnosing mental health issues, managing depression, evaluating suicide risk, and supporting education in the field. However, our review also points out their limitations, such as difficulties with complex cases and potential underestimation of suicide risks.

**Conclusion:** Early research in psychiatry reveals LLMs’ versatile applications, from diagnostic support to educational roles. Given the rapid pace of advancement, future investigations are poised to explore the extent to which these models might redefine traditional roles in mental health care.

## Introduction

The integration of artificial intelligence (AI) into various healthcare sectors has brought transformative changes (1–3). Currently, Large Language Models (LLMs) like Chat Generative Pre-trained Transformer (ChatGPT) are at the forefront (2,4,5).

Advanced LLMs, such as GPT-4 and Claude Opus, possess an uncanny ability to understand and generate human-like text. This capacity indicates their potential to act as intermediaries between AI functionalities and the complexities of human cognition.

Unlike their broader application in healthcare, LLMs in psychiatry address unique challenges such as the need for personalized mental health interventions and the management of complex mental disorders (4,6). Their capacity for human-like language generation and interaction is not just a technological advancement; it’s a critical tool in bridging the treatment gap in mental health, especially in under-resourced areas (4,6–8).

In psychiatry, LLMs like ChatGPT can provide accessible mental health services, breaking down geographical, financial, or temporal barriers, which are particularly pronounced in mental health care (4,9). For instance, ChatGPT can support therapists by offering tailored assistance during various treatment phases, from initial assessment to post-treatment recovery (10–12). This includes aiding in symptom management and encouraging healthy lifestyle changes pertinent to psychiatric care (10–14).

ChatGPT’s ability to provide preliminary mental health assessments and psychotherapeutic support is a notable advancement (15,16). It can engage in meaningful conversations, offering companionship and empathetic responses, tailored to individual mental health needs (6,10,11,13), a component that is essential in psychiatric therapy (17).

Despite these capabilities, currently, LLMs do not replace human therapists (18–20). Rather, the technology supplements existing care, enhancing the overall treatment process while acknowledging the value of human clinical judgment and therapeutic relationships (4,8,13).

As LLM technology advances rapidly, it holds the potential to alter traditional mental health care paradigms. This review aims to assess their current role within psychiatry research.

## Methods

### Search Strategy

The review was registered with the International Prospective Register of Systematic Reviews - PROSPERO (Registration code: CRD42024524035) We adhered to the Preferred Reporting Items for Systematic Reviews and Meta-Analyses (PRISMA) guidelines (21,22).

A systematic search was conducted across key databases: PubMed, Embase, Web of Science, and Scopus, from December 2022 up until March 2024. We chose December 2022, as it was the date of introduction of chatGPT. We complemented the search via reference screening for any additional papers. We chose PubMed, Embase, Web of Science, and Scopus for their comprehensive coverage of medical and psychiatric literature.

Our search strategy combined specific keywords related to LLM, including ’ChatGPT,’ ’Artificial Intelligence,’ ’Natural Language Processing,’ and ’Large Language Models,’ with psychiatric terminology such as ’Psychiatry’ and ’Mental Health.’ Additionally, to refine our search, we incorporated keywords for the most relevant psychiatric diseases. These included terms like ’Depression,’ ’Anxiety Disorders,’ ’Bipolar Disorder,’ ’Schizophrenia,’ and others pertinent to our study’s scope.

Specific search strings for each database are detailed in the supplementary materials.

### Study Selection

The selection of studies was rigorously conducted by two independent reviewers, MO and EK. Inclusion criteria were set to original research articles that specifically examined the application of LLMs in psychiatric settings.

Eligible studies were required to present measurable outcomes related to psychiatric care, such as patient engagement, diagnostic accuracy, treatment adherence, or clinician efficiency.

We excluded review articles, case reports, conference abstracts without full texts, editorials, preprints, and studies not written in English.

MO and EK systematically evaluated each article against these criteria. In cases of disagreement or uncertainty regarding the eligibility of a particular study, the matter was resolved through discussion and, if necessary, consultation with additional researchers in our team to reach a consensus.

### Data Extraction

Data extraction was performed by two independent reviewers, MO and EK, using a structured template. Key information extracted included the study’s title, authors, publication year, study design, psychiatric condition or setting, the role of the LLM model, sample size, findings related to the effectiveness and impact of the model, and any noted conclusions and implications. In cases of discrepancy during the extraction process, issues were resolved through discussion and consultation with other researchers involved in the study.

#### Risk of Bias

In our systematic review, we opted for a detailed approach instead of a standard risk of bias assessment, given the unique and diverse nature of the studies included. Each study is presented in a table highlighting its design and essential variables **(Table 1)**.

**Table 1:** Summary of the included papers.

| Author | Year | Model | Application | Main Results |
| --- | --- | --- | --- | --- |
| Liyanage et al.(36) | 2023 | GPT-3.5 | Data augmentation for wellness dimension classification | ChatGPT models effectively augmented Reddit post data, significantly improving classification performance for wellness dimensions. |
| Hwang et al.(37) | 2024 | GPT-4 | Psychodynamic formulations in psychiatry | GPT-4 successfully created relevant and accurate psychodynamic formulations based on patient history. |
| Parker et al.(35) | 2023 | GPT-3 | Information on bipolar disorder | GPT-3 provided basic material on bipolar disorders and creative song generation, but lacked depth for scientific review. |
| Heston T.F. et al.(34) | 2023 | GPT-3.5 | Simulating depression scenarios | ChatGPT-3.5 conversational agents recommended human support at critical points, highlighting the need for AI safety in mental health. |
| D'Souza et al.(33) | 2023 | GPT-3.5 | Responding to psychiatric case vignettes | ChatGPT 3.5 showed high competence in handling psychiatric case vignettes, with strong diagnostic and management strategy generation. |
| Levkovich et al.(31) | 2023 | GPT-3.5, GPT-4 | Depression diagnosis and treatment | ChatGPT aligned with guidelines for depression management, contrasting with primary care physicians and showing no gender or socioeconomic biases. |
| Mazumdar et al.(32) | 2023 | GPT-3, BERT-large, MentalBERT, ClinicBERT, and PsychBERT | Mental health disorder classification and explanation | GPT-3 outperformed other models in classifying mental health disorders and generating explanations, showing promise for AI-IoMT deployment. |
| Sezgin et al.(38) | 2023 | GPT-4, LaMDA (using Bard) | Responding to postpartum depression questions | GPT-4 provided more clinically accurate responses to postpartum depression questions, surpassing other models and Google Search. |
| Elyoseph et al.(30) | 2023 | GPT-3.5 | Emotional awareness evaluations | ChatGPT showed higher emotional awareness compared to the general population and improved over time. |
| Elyoseph et al.(29) | 2023 | GPT-3.5 | Suicide risk assessment | ChatGPT underestimated suicide risks compared to mental health professionals, indicating the need for human judgment in complex assessments. |
| Levkovich et al.(28) | 2023 | GPT-3.5, GPT-4 | Suicide risk assessment vignettes | GPT-4's evaluations of suicide risk were similar to mental health professionals, though with some overestimations and underestimations. |
| Dergaa et al.(27) | 2024 | GPT-3.5 | Mental health assessment and interventions | ChatGPT showed limitations in complex medical scenarios, underlining its unpreparedness for standalone use in mental health practice. |
| Spallek et al.(26) | 2023 | GPT-4 | Mental health and substance use education | GPT-4's outputs were substandard compared to expert materials in terms of depth and adherence to communication guidelines. |
| Hadar-Shoval D et al.(25) | 2023 | GPT-3.5 | Responses for BPD and SPD scenarios | ChatGPT effectively differentiated emotional responses in BPD and SPD scenarios, showing tailored mentalizing abilities. |
| Elyoseph et al.(24) | 2024 | GPT-3.5, GPT-4 | Prognosis in depression | ChatGPT-3.5 showed a more pessimistic prognosis in depression compared to other LLMs and mental health professionals. |
| Li et al.(23) | 2024 | GPT-4, Bard and Llama-2 | Psychiatric licensing examination and diagnostics | GPT-4 outperformed other models in psychiatric diagnostics, closely matching the capabilities of human psychiatrists. |

Abbreviations: BPD: Borderline Personality Disorder | SPD: Schizoid Personality Disorder | GPT: Generative Pre-trained Transformer | AI: Artificial Intelligence | NLP: Natural Language Processing | EDA: Easy Data Augmentation | BT: Back Translation | PHQ-9: Patient Health Questionnaire-9 | LEAS: Levels of Emotional Awareness Scale | AI-IoMT: Artificial Intelligence-Internet of Medical Things | GRADE: Grading of Recommendations, Assessment, Development, and Evaluations.

A second table catalogs the inherent limitations of each study, providing a transparent overview of potential biases and impacts on the results **(Table 2)**.

**Table 2:**
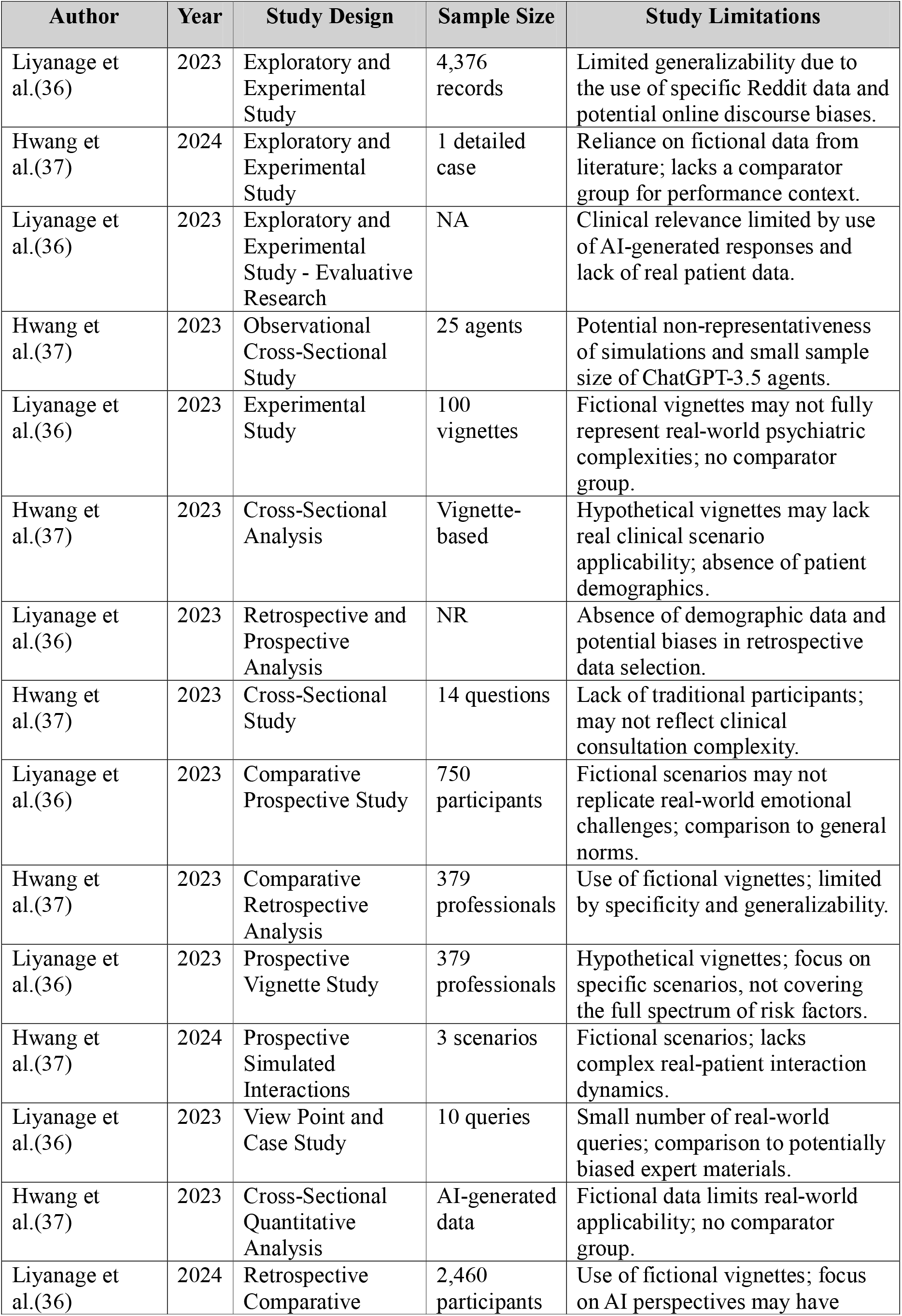
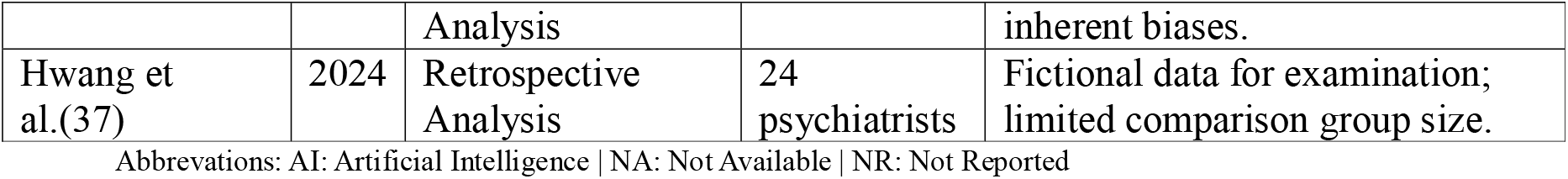
Summary of the studies designs and limitations.

Additionally, a figure illustrates the quartiles and SCImago Journal Rank scores of the journals where these studies were published, offering insight into their academic significance **(Figure 1)**. This method ensures a clear, concise evaluation of the varied included papers.

**Figure 1:**
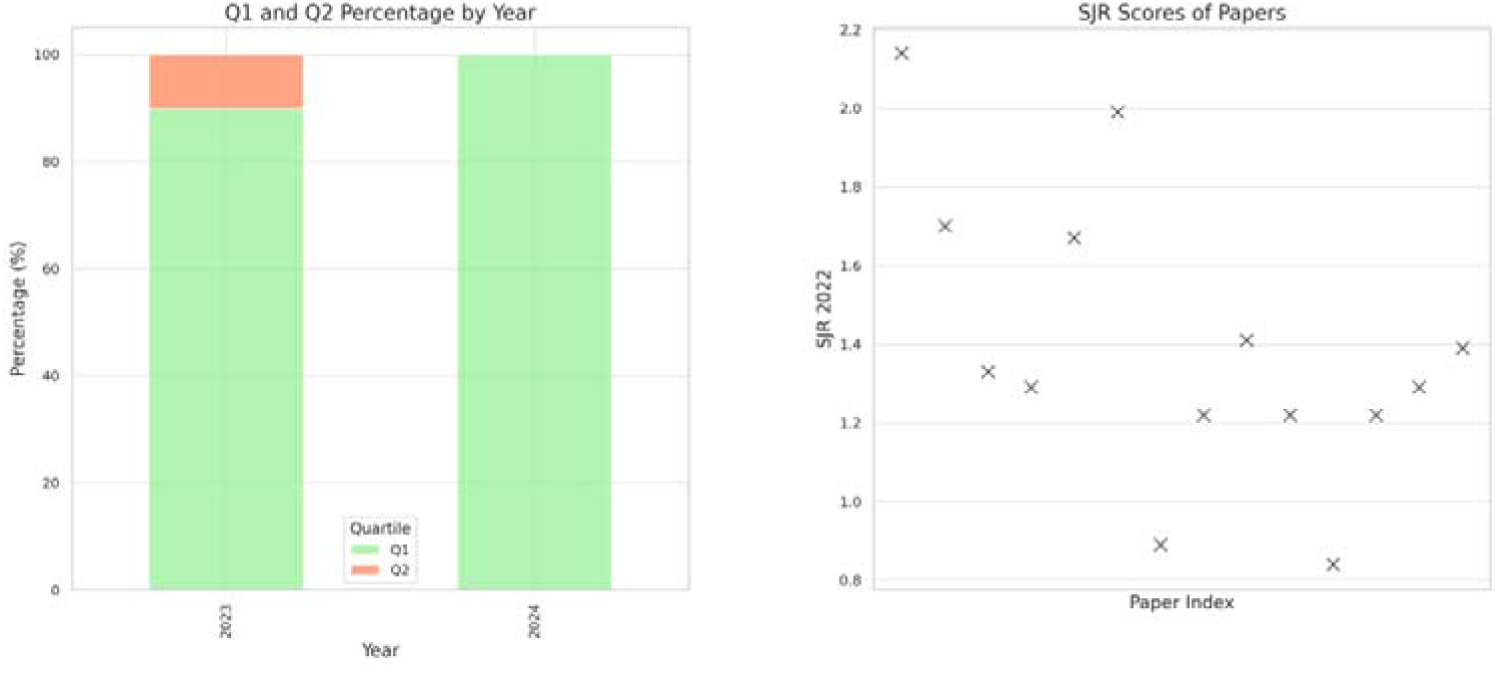
SJR scores and journal quartiles of the included studies.

## Results

### Search Results and Study Selection

Our systematic search across PubMed, Embase, Web of Science, and Scopus yielded a total of 771 papers. The breakdown of the initial results was as follows: PubMed (186), Scopus (290), Embase (133), and Web of Science (162). After applying automated filters to exclude review articles, case reports, and other non-relevant document types, 454 articles remained. The removal of duplicates further reduced the pool to 288 articles.

Subsequent screening based on titles and abstracts led to the exclusion of 255 papers, primarily due to their irrelevance or lack of discussion on LLMs, leaving 33 articles for full-text evaluation. Upon detailed examination, 16 studies were found to meet our inclusion criteria and were thus selected for the final review (23–38). The process of study selection and the results at each stage are comprehensively illustrated in **Figure 2**, the PRISMA flowchart.

**Figure 2:**
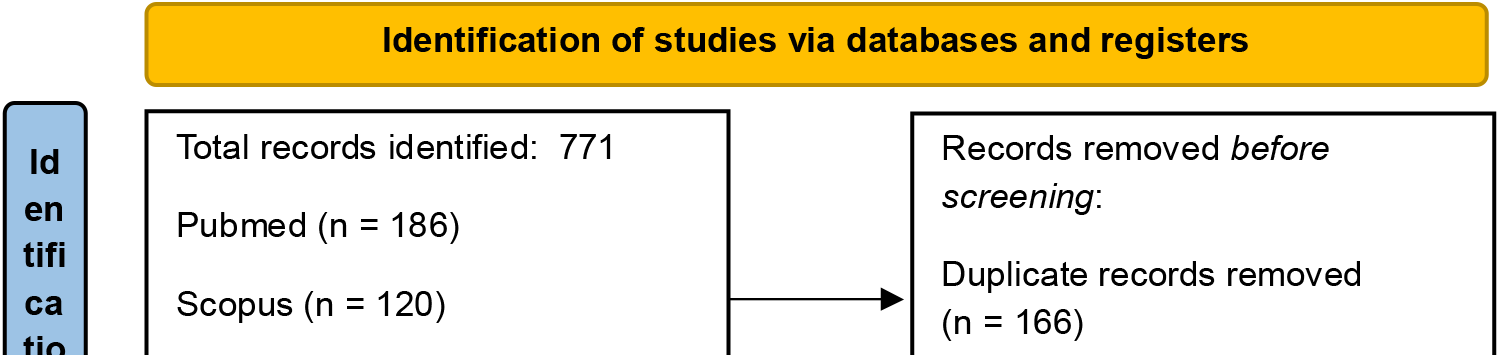
PRISMA flowchart.

### Overview of the Included Studies

Most of the studies in our review were published in Q1 journals, indicating a high level of influence in the field (**Figure 1**). The studies varied in their approach, using data ranging from real patient interactions on online platforms to simulated scenarios, and in scale, from individual case studies to large datasets.

Most research focused on various versions of ChatGPT, including ChatGPT-3.5 and ChatGPT-4, with some comparing its performance to traditional methods or other LLMs. The applications of LLMs in these studies were diverse, covering aspects like mental health screening augmentation on social media, generating psychodynamic formulations, and assessing risks in psychiatric conditions.

In highlighting key studies, Liyanage et al. found that ChatGPT was effective in enhancing Reddit post analysis for wellness classification (36). Levkovich et al. observed ChatGPT’s unbiased approach in depression diagnosis, contrasting with biases noted in primary care physicians’ methods, especially due to gender and socioeconomic status (31). Additionally, Li et al. demonstrated GPT-4’s proficiency in psychiatric diagnostics, uniquely passing the Taiwanese Psychiatric Licensing Examination and paralleling experienced psychiatrists’ diagnostic abilities (23).

### LLMs’ applications and limitations in mental health

We categorized the applications of LLM in the included studies into three main themes to provide a synthesized and comprehensive overview:

<u>Applications (**Table 1, Figure 3**)</u>

**1. Clinical Reasoning:**
  - **Hwang et al.** demonstrated ChatGPT’s ability in generating psychodynamic formulations, indicating potential in clinical psychiatry with statistical significance (Kendall’s W = 0.728, p = 0.012) (37).
  - **Levkovich et al.** (**2023**) compared ChatGPT’s recommendations for depression treatment against primary care physicians, finding ChatGPT more aligned with accepted guidelines, particularly for mild depression (31).
  - **Levkovich et al. and Elyoseph et al.** (**2023**) assessed ChatGPT’s performance in assessing suicide risk. The study by Levkovich et al. highlighted that ChatGPT tended to underestimate risks when compared to mental health professionals, particularly in scenarios with high perceived burdensomeness and feeling of thwarted belongingness (28). The study be Elyoseph, found that while GPT-4’s evaluations were similar to mental health professionals, ChatGPT-3.5 often underestimated suicide risk (29).
  - **D’Souza et al.** evaluated ChatGPT’s response to psychiatric case vignettes, where it received high ratings, especially in generating management strategies for conditions like anxiety and depression (33).
  - **Li et al.** demonstrated ChatGPT GPT-4’s capabilities in the Taiwanese Psychiatric Licensing Examination and psychiatric diagnostics, closely approximating the performance of experienced psychiatrists (23). ChatGPT outperformed the two LLMs, Bard and Llama-2.
  - **Heston T.F. et al.** Evaluated ChatGPT-3.5’s responses in depression simulations. AI typically recommended human support at moderate depression levels (PHQ-9 score of 12) and insisted on human intervention at severe levels (score of 25) (34).
  - **Dergaa et al.** critically assessed ChatGPT’s effectiveness in mental health assessments, particularly highlighting its inadequacy in dealing with complex situations, such as nuanced cases of postpartum depression requiring detailed clinical judgment, suggesting limitations in its current readiness for broader clinical use (27).
  - **Elyoseph et al.** (**2024**) provided a comparative analysis of depression prognosis from the perspectives of AI models, mental health professionals, and the general public. The study revealed notable differences in long-term outcome predictions. AI models, including ChatGPT, showed variability in prognostic outlooks, with ChatGPT-3.5 often presenting a more pessimistic view compared to other AI models and human evaluation (24).
  - **Elyoseph et al.** (**2023**) investigated ChatGPT’s emotional awareness using the Levels of Emotional Awareness Scale (LEAS). ChatGPT scored significantly higher than the general population, indicating a high level of emotional understanding (30).
**2. Social Media applications:**
  - **Liyanage et al.** used ChatGPT models to augment data from Reddit posts, enhancing classifier performance in identifying Wellness Dimensions. This resulted in improvements in the F-score of up to 13.11% (36).
  - **Mazumdar et al.** applied GPT-3 in classifying mental health disorders from Reddit data, achieving an accuracy of around 87% and demonstrating its effectiveness in explanation generation (32). GPT-3 demonstrated superior performance in classifying mental health disorders and generating explanations, outperforming traditional models like LIME and SHAP.

**Figure 3:**
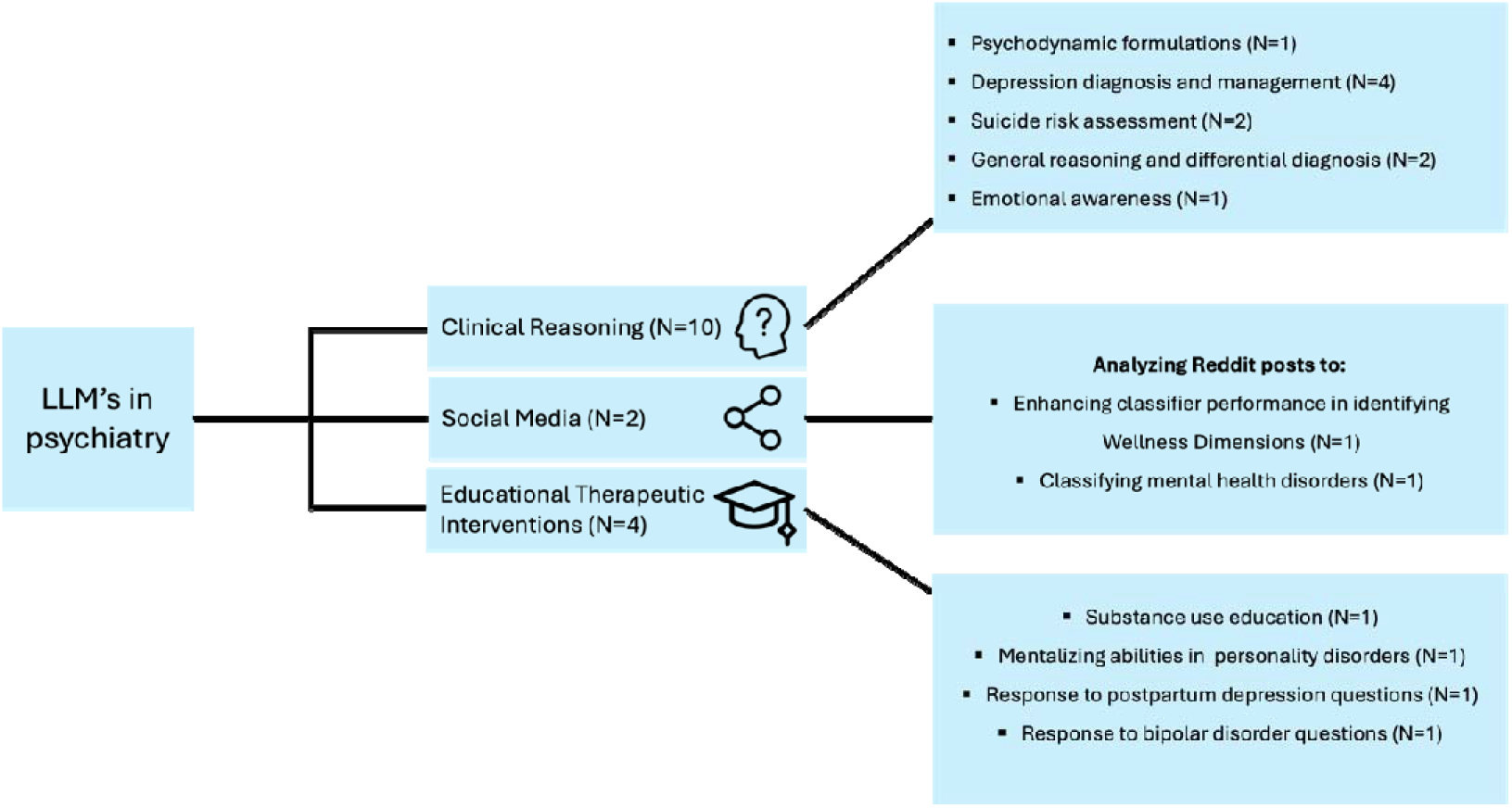
Applications and Evaluations of LLMs in Diverse Domains of Psychiatry.

Figure 4 presents the different types of data inputs for GPT in the current applications for the field of psychiatry.

**3. Educational Therapeutic Interventions:**
  - **Spallek et al.** examined ChatGPT’s application in mental health and substance use education, finding its outputs to be substandard compared to expert materials. However, when prompts where carefully engineered, the outputs were better aligned with communication guidelines (26).
  - **Hadar-Shoval D et al.**, explored ChatGPT’s ability to understand mental state in personality disorders (25), and **Sezgin et al.** (38), assessed responses to postpartum depression questions Both studies reflect ChatGPT’s utility both as an educational resource and for offering preliminary therapeutic advice. **Sezgin et al.** (38) showed that GPT-4 demonstrated generally higher quality, more clinically accurate responses compared to Bard and Google Search.
  - **Parker et al.** ChatGPT-3.5 was used to respond to clinically relevant questions about bipolar disorder and to generate songs related to bipolar disorder, testing both its factual knowledge and creativity. The study highlighted its utility in providing basic information, but also its limitations in citing current, accurate references (35).

**Figure 4:**
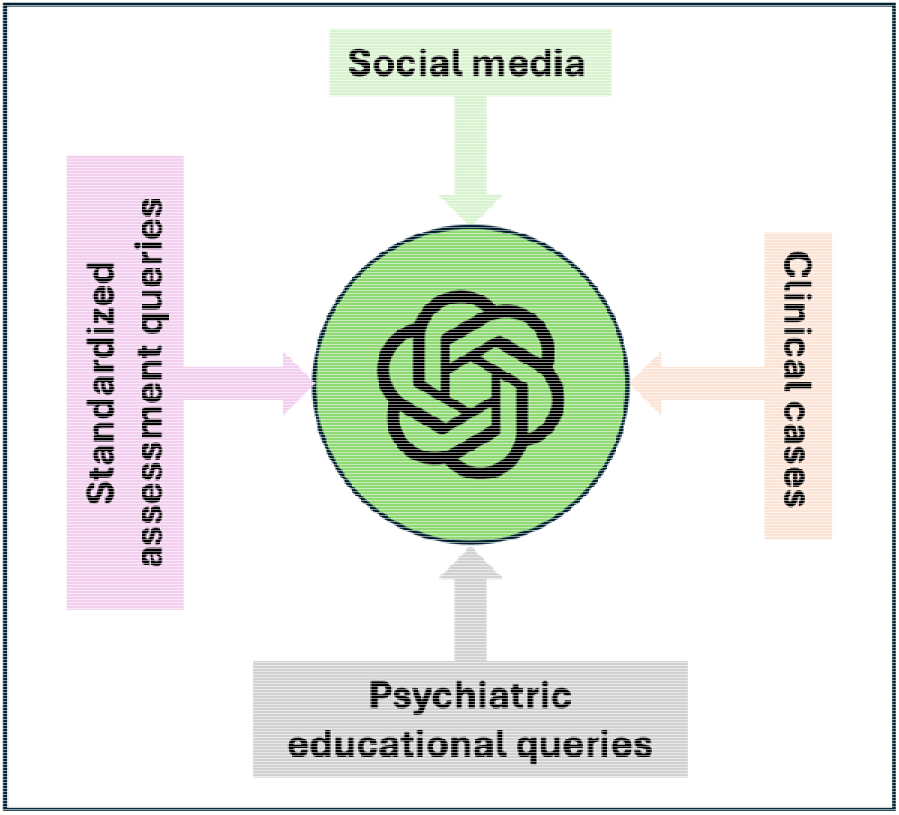
Data Input Spectrum for GPT in Psychiatric Applications.

Additional data in the supplementary material includes demographics, journals of the included papers, SCImago Journal Rank (SJR) 2022 data for these publications, and specific performance metrics for the models across various applications, detailed in **S1-S2 tables** in the supplementary material.

#### Safety and Limitations

Concerns regarding safety and limitations in LLMs clinical applications emerge as critical themes. For instance, Heston T.F. et al. observed that ChatGPT-3.5 recommended human support at moderate depression levels but only insisted on human intervention at severe levels, underscoring the need for cautious application in high-risk scenarios (34).

Elyoseph et al. highlighted that ChatGPT consistently underestimated suicide risks compared to mental health professionals, especially in scenarios with high perceived burdensomeness and thwarted belongingness (29).

Dergaa et al. concluded that ChatGPT, as of July 2023, was not ready for mental health assessment and intervention roles, showing limitations in complex case management (27). This suggest that while ChatGPT shows promise, it is not without significant risks and limitations, particularly in handling complex and sensitive mental health scenarios (**Table 2**, Figure 5).

**Figure 5:**
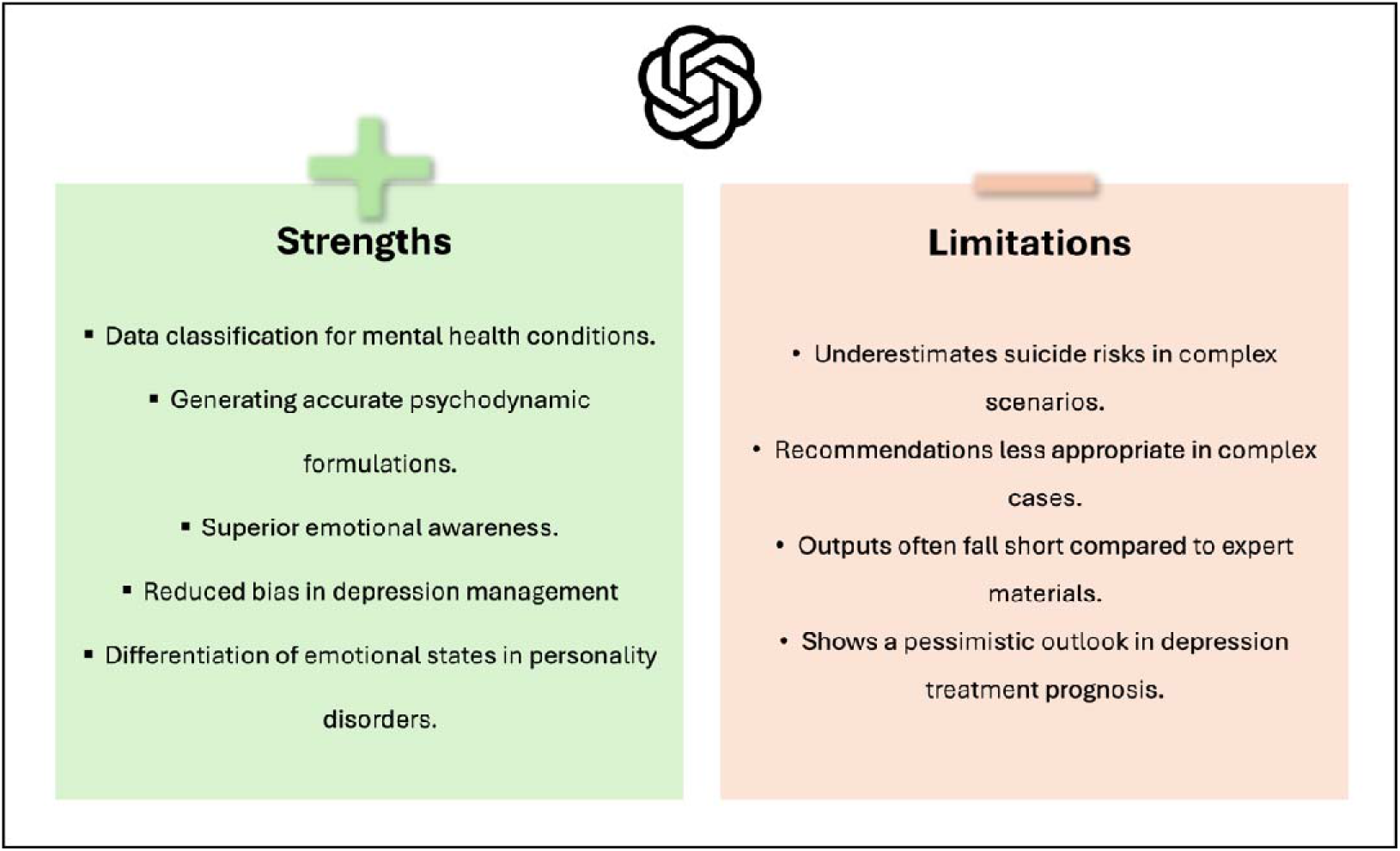
Strengths and limitations of GPT’s current applications in psychiatry practice.

## Discussion

Our findings demonstrate LLMs, especially ChatGPT and GPT-4, potential as a valuable tool in psychiatry, offering diverse applications from clinical support to educational roles. Studies like Liyanage et al. and Mazumdar et al. showcased its efficacy in data augmentation and mental health disorder classification (32,36). Others, such as Hwang et al. and Levkovich et al. (2023), highlighted its capabilities in clinical settings, including diagnosis and risk assessment (31,37). Overall, GPT emerged as the most used and studied LLM in the field of psychiatry.

However, concerns about its limitations and safety in clinical scenarios were evident, as seen in studies by Elyoseph et al. (2023) and Dergaa et al., indicating that while ChatGPT holds promise, its integration into clinical psychiatry must be approached with caution (27,29).

The potential and efficacy of AI, particularly LLMs, in psychiatry are highlighted by our review, showing its capability to streamline care, lower barriers, and reduce costs in mental health services (39). Studies like Liyanage et al. and Hwang et al. illustrate ChatGPT’s diverse applications, from clinical data analysis to formulating psychodynamic profiles, which contribute to a more efficient, accessible, and versatile approach in mental healthcare (19,20,36,37). Moreover, in light of the COVID-19 pandemic’s impact on mental health and the growing demand for digital interventions, Mitsea et al.’s research highlights the significant role of AI in enhancing digitally assisted mindfulness training for self-regulation and mental well-being, further enriching the scope of AI applications in mental healthcare (40).

When compared with humans and other LLMs, ChatGPT consistently adheres to clinical guidelines, as shown in Levkovich et al.’s study (31). Furthermore, ChatGPT often surpasses other models in tasks like psychiatric diagnostics, as demonstrated by Li et al (23).

These studies collectively underscore the utility of current LLMs, particularly ChatGPT and GPT-4, in psychiatry, demonstrating promise across various domains. While serving as a complementary tool to human expertise, especially in complex psychiatric scenarios (4,6,11,20), LLMs are poised for deeper integration into mental health care. This evolution is propelled by rapid technological advancements and significant financial investments since the watershed moment of ChatGPT introduction, late 2022. Future research should closely monitor this integration, exploring how LLMs not only supplement but also augment human expertise in psychiatry.

Our review has limitations. The absence of a formal risk of bias assessment, due to the unique nature of the included studies, is a notable drawback. Additionally, the reliance on studies that did not use real patient data as well as the heterogeneity in study designs could affect the generalizability of our findings. Moreover, the diversity of methods and tasks in the included studies prohibited us from performing a meta-analysis. It should also be mentioned that all studies were retrospective in nature. Future directions should include prospective, real-world evidence studies, that could cement the utility of LLM in the psychiatry field.

In conclusion,

Early research in psychiatry reveals LLMs’ versatile applications, from diagnostic support to educational roles. Given the rapid pace of advancement, future investigations are poised to explore the extent to which these models might redefine traditional roles in mental health care.

## Data Availability

All data produced in the present work are contained in the manuscript

## Acknowledgements

The authors wish to thank the research and administrative staff at their respective institutions for their support and contributions to this study.

## Author Contributions

Mahmud Omar (MO): Conceptualization, Methodology, Screening and Data Extraction, Writing - Original Draft Preparation.

Shelly Soffer: Data Collection, Writing - Review & Editing.

Alexander W Charney: Supervision, Validation, Writing - Review & Editing.

Isotta Landi: Data Interpretation, Writing - Review & Editing.

Girish N Nadkarni: Project Administration, Resources, Writing - Review & Editing.

Eyal Klang (EK): Data Curation, Screening and Data Extraction, Writing - Review & Editing.

## Ethical Approval

Not applicable for this systematic review.

## Consent for Publication

All authors have read and agreed to the published version of the manuscript.

